# Conservative treatment of partial-thickness rotator cuff tears and tendinopathy with platelet-rich plasma: a systematic review and meta-analysis

**DOI:** 10.1101/2020.12.19.20248575

**Authors:** Xiao-Na Xiang, Jie Deng, Yan Liu, Xi Yu, Biao Cheng, Hong-Chen He

**Author notes:** These authors contributed equally to this manuscript (Xiao-Na Xiang, Jie Deng).

## Abstract

**Purpose:** To assess the effect of platelet-rich plasma (PRP) as a conservative therapy on individuals with partial-thickness rotator cuff tears (PTRCs) or tendinopathy in terms of pain, and function.

**Methods:** A systematic review and meta-analysis of randomized controlled trials were conducted. Short-term (6±1 months) and long-term (≥1 year) outcomes were analysed, including the visual analogue scale (VAS), Constant-Murley score (CMS), Shoulder Pain and Disability Index (SPADI), as well as American Shoulder and Elbow Surgeons (ASES) score. The weighted mean difference (MD) with 95% confidence interval (CI) was used.

**Results:** Ten studies were eligible in this review, nine studies with 561 patients were included in this meta-analysis. The meta-analysis showed statistically significant differences in the decrease in short-term VAS (MD=-1.56; 95% CI -2.82 to -0.30), and increase in CMS (MD=16.48; 95% CI 12.57 to 20.40), and SPADI (MD=-18.78 95% CI -36.55 to -1.02). Nonetheless, no long-term effect was observed on pain and function, except CMS (MD=24.30). The results of mean important differences (MIDs) reached the minimal clinically important differences, except ASES. For subgroup analysis, short-term VAS scores were decreased in PRP-treated patients with double centrifugation (MD=-1.99), single injection (MD=-0.71) and post-injection rehabilitation (MD=-1.59).

**Conclusion:** PRP-treated patients with PTRCs and rotator cuff tendinopathy demonstrated improvements in pain and function, although the effect may not last for a long time. Overall, our results suggest that PRP may have positive clinical outcomes, but limited data, and study heterogeneity hinder firm conclusions.

## 1. Introduction

Partial-thickness rotator cuff tears (PTRCs) and rotator cuff tendinopathy commonly affected overhead athletes,^1^ such as handball players^2^ and pitchers,^3^ which leads to sport performance decline. Rotator cuff tendinopathy, as an overuse disorder,^4^ is the most common cause of shoulder pain,^5^ which has been associated with an increased risk for rotator cuff tears.^6^ Although PTRCs may cause minimal symptoms instantly,^7^ concomitant pathologic physical condition such as scapula instability, dyskinesis, or tightness may cause pain and muscle weakness.^8^ PTRCs and tendinopathy are initially treated conservatively^9^ with sodium hyaluronate^10^ and corticosteroid,^11^ saline, lidocaine, and physiotherapy,^12^ etc. Additionally, the limited ability of the tendon to heal has led to increased interest in platelet-rich plasma (PRP).

PRP is an autologous product, which is separated from the fresh whole blood after centrifugation.^13, 14^ It contains numerous growth factors (such as platelet-derived growth factor AB, transforming growth factor β-1, and vascular endothelial growth factor) which promote tissue repair.^15^ However, the clinical effect of PRP is numerous, due to its difference in component, biologic characteristic, post-injection rehabilitation, and PRP preparation, especially the inconsistent PRP parameters (preparation kit, platelet concentration and the presence of cells, etc).

Recently, numerous studies have researched the healing effect of PRP injection in rotator cuff disease. A systematic review of rotator cuff disease has reported that PRP injections may not be beneficial at short-term follow-up.^16^ Moreover, in the latest guideline by the American Academy of Orthopaedic Surgeons (AAOS),^17^ the routine use of PRP in the non-operative management of PTRCs is not recommended by limited quality and number of evidences. Nonetheless, several trials have reported the positive effects of PRP injection for PTRCs or rotator cuff tendinopathy,^18, 19^ while no meta-analysis study yet. To our’ knowledge, this is the first meta-analysis study of randomized controlled clinical trials aiming at the effectiveness of PRP in PTRCs and rotator cuff tendinopathy. Furthermore, the minimal clinically important differences (MCIDs) are important considerations in making clinical conclusion and are discussed in this study.^20, 21^

## 2. Materials and Methods

The review protocol was registered with PROSPERO (CRD42020165330) and was performed according to the PRISMA recommendations (Preferred Reporting Items for Systematic Reviews and Meta-analyses).^22^

### 2.1 Search Strategy

Electronic databases were searched from inception to September 2020 for randomized controlled trials (RCTs) that involved Embase (via OVID platform), MEDLINE (via OVID platform), CENTRAL (via the Cochrane Library), Web of Science, CINAHL (via EBSCOhost) and the Physiotherapy Evidence Database. Two authors (X.N.X. and J.D.) independently performed the initial screening and study selection. Any disagreements were resolved by a discussion under the guidance of a third reviewer (Y.L.). The key search terms included “rotator cuff injuries”, “rotator cuff tendinopathy”, “PRP or platelet rich plasma”, “randomized controlled trial” and “human”. We developed a search strategy via OvidSP based on the aforementioned. The full search strategy is listed in Supplementary Material. The specialist register GreyNet (http://www.greynet.org/) for grey literature was also searched. The reference lists of potentially relevant articles were also hand-searched.

### 2.2 Study selection and inclusion criteria

The inclusion criteria were as follows: (1) patients diagnosed with PTRCs or rotator cuff tendinopathy by MRI or ultrasound over 2 months; (2) patients aged 18 years or older; (3) patients in the intervention group received PRP; (4) quantifiable outcomes were reported; (5) RCT design. Exclusion criteria were as followed: (1) patients diagnosed with full-thickness rotator cuff tears, rheumatoid arthritis, adhesive capsulitis, calcific rotator disease; (2) patients had a history of rotator cuff repair or injection; (3) case reports, letters, comments, trial protocols, editorials, reviews or practice guidelines; (4) studies were not written in English.

### 2.3 Data extraction and analysis

Outcome measures: the primary outcome was pain assessed by visual analogue scale (VAS) score; the secondary outcome was function that consisted of the Constant-Murley Score (CMS), Disabilities of the Arm, Shoulder Pain and Disability Index (SPADI), and American Shoulder and Elbow Surgeons (ASES).

Two independent authors (X.N.X. and J.D.) extracted data with standardized data forms. The following data were extracted: first author’s name, publication year, percentage of female patients, mean age, outcome measurements, details of PRP treatment and duration of follow-up. The means and standard deviations (SD) of continuous outcomes were recorded. All other data were recorded as median with corresponding. The VAS score (reported on 0 -100 scales) was converted into 0-10 scales. The results of the VAS score, CMS, SPADI, and ASES were extracted and categorized as follows: baseline, short-term (6±1 months) follow-up, and long-term (≥1-year) follow-up. The data of diagram was extracted by Engauge Digitizer (version 3.0), if no original data available in the study. If SDs were missing for continuous data, other statistics (for example: 95% confidence interval, standard errors, *T* values, *F* values, and *P* values) were used for the calculation of standard deviation via the calculator tool from Review Manager, version 5.3 (Nordic Cochrane Centre, Cochrane Collaboration). Disagreements between the two reviewers were resolved by consensus, and if necessary, by consultation with a third reviewer (Y.L.).

Data analysis was performed with Review Manager. A *P* value <.05 was considered statistically significant. For the continuous variable, the weighted mean difference (MD) with 95% confidence interval (CI) was used. the unit of measurement was the same across studies for the specified outcomes and the MD was not standardized. Heterogeneity was assessed using Cochrane *Q* statistic (significance level at *P* value< .05) and quantified with *I*^*2*^ (significance level at *I*^*2*^>50%).^23, 24^ Random effects were used if the *Q* or *I*^*2*^ value was statistically significant or a small number of studies was analysed. Otherwise, fixed effects were used. A sensitivity analysis was performed for primary and secondary outcomes by excluding studies with high risk of bias^25^. The subgroup analysis was reported based on several variables of interest (PRP centrifugation approach, number of injections, and the existence of post-injection rehabilitation).

According to previous studies the values of MCID were reported as 1.0 (VAS),^26^ 5.5(CMS), 14.88 (SPADI), and 11.1 (ASES),^27, 28^ respectively. In this study, the minimal important differences (MIDs) between two groups calculated by anchor-based methods^29^ were compared with those MCIDs. Therefore, clinical improvements were deemed meaningful when the MIDs calculated were beyond MCIDs.

### 2.4 Assessment of risk of bias in include studies

To assess bias with the Cochrane Collaboration’s risk of bias tool, two reviewers (X.N.X. and J.D.) independently assessed each of following domains: selection bias, performance bias, detection bias, attrition bias, reporting bias and other sources of bias. Each component was recorded as low, unclear, or high risk of bias. Studies were considered as low risk of bias only when a “+” was recorded for all items; a high risk of bias was considered if studies scored “-” or “?” more than two items. Others were scored as moderate risk of bias.^30, 31^ Any disagreements were resolved through discussion or by referral to a third author (Y.L.).

## 3. Results

### 3.1 Study characteristics

Studies retrieved and excluded in each screening phase appear in Figure 1. A total of ten (601 patients) level 1 studies^5, 32-40^ was deemed eligible for systematic review and 9 studies (561 patients) were included in the meta-analysis. One study did not include the primary outcome and was removed.^39^ The mean age of the included participants was 49 years (range, 40-64 years); 55% of the participants were female. One study^37^ was extracted from figure to number via Engauge Digitizer.

**Figure 1.**
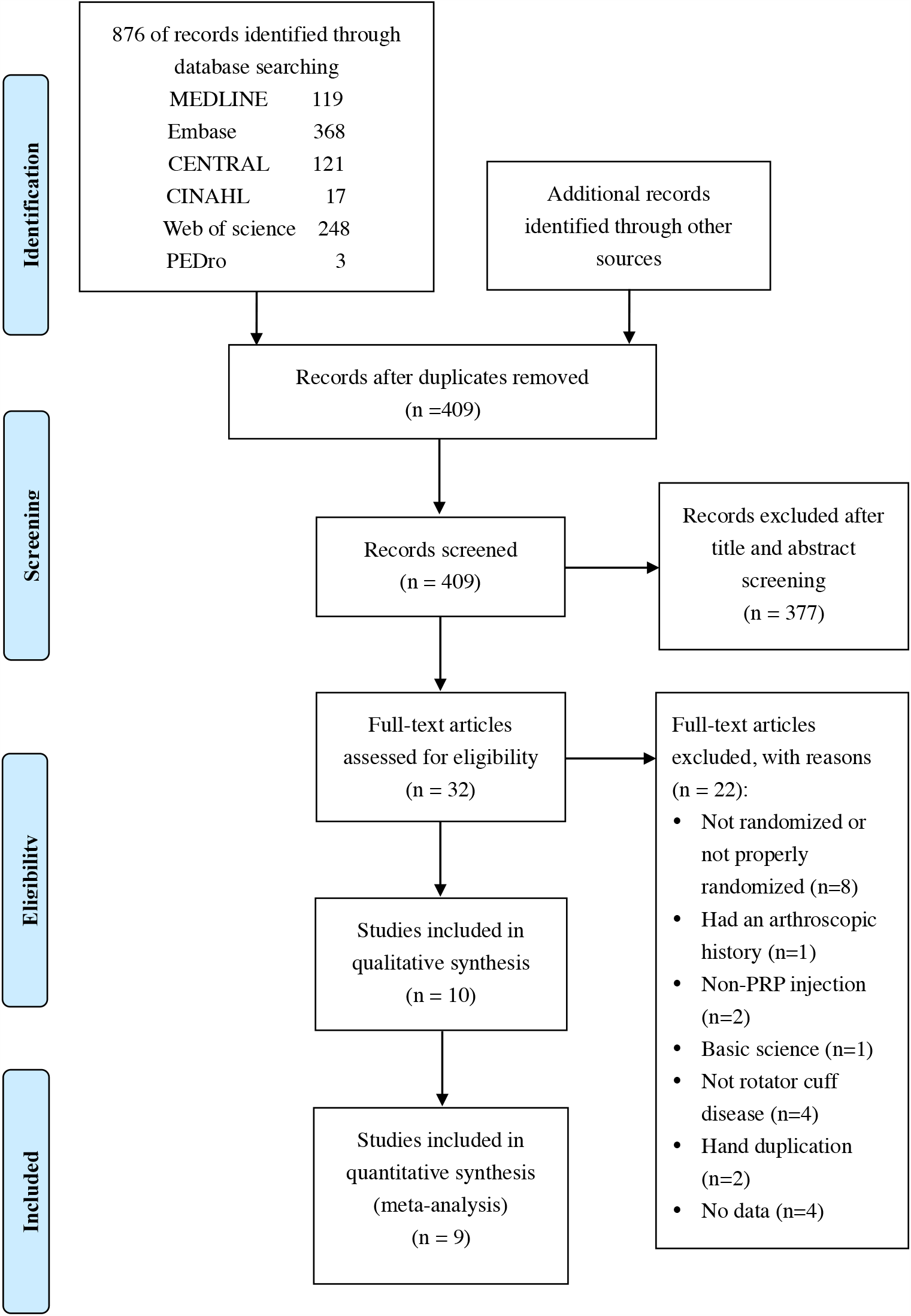
the graph shows a PRISMA (Preferred Reporting Items for Systematic Reviews and Meta-Analyses) flow diagram of the study selection process.

PRP treatment characteristics varied widely among the included studies, though all injections were under ultrasound guidance. For example, PRP parameters included the preparation kit used in five trials,^5, 33, 38-40^ the presence of leukocyte in three trials,^33-35^ platelet concentration in three trials^5, 32, 34^ and activation agents in seven trials.^5, 32-37^ After injection, seven studies followed standard rehabilitation programs for rotator cuff tendinopathy or partial-thickness tear.^5, 32, 33, 37-40^ Two RCTs^33, 34^ avoided medicine and physical therapy for several months after injections. Included study characteristics are provided in Table 1.

**Table 1.**
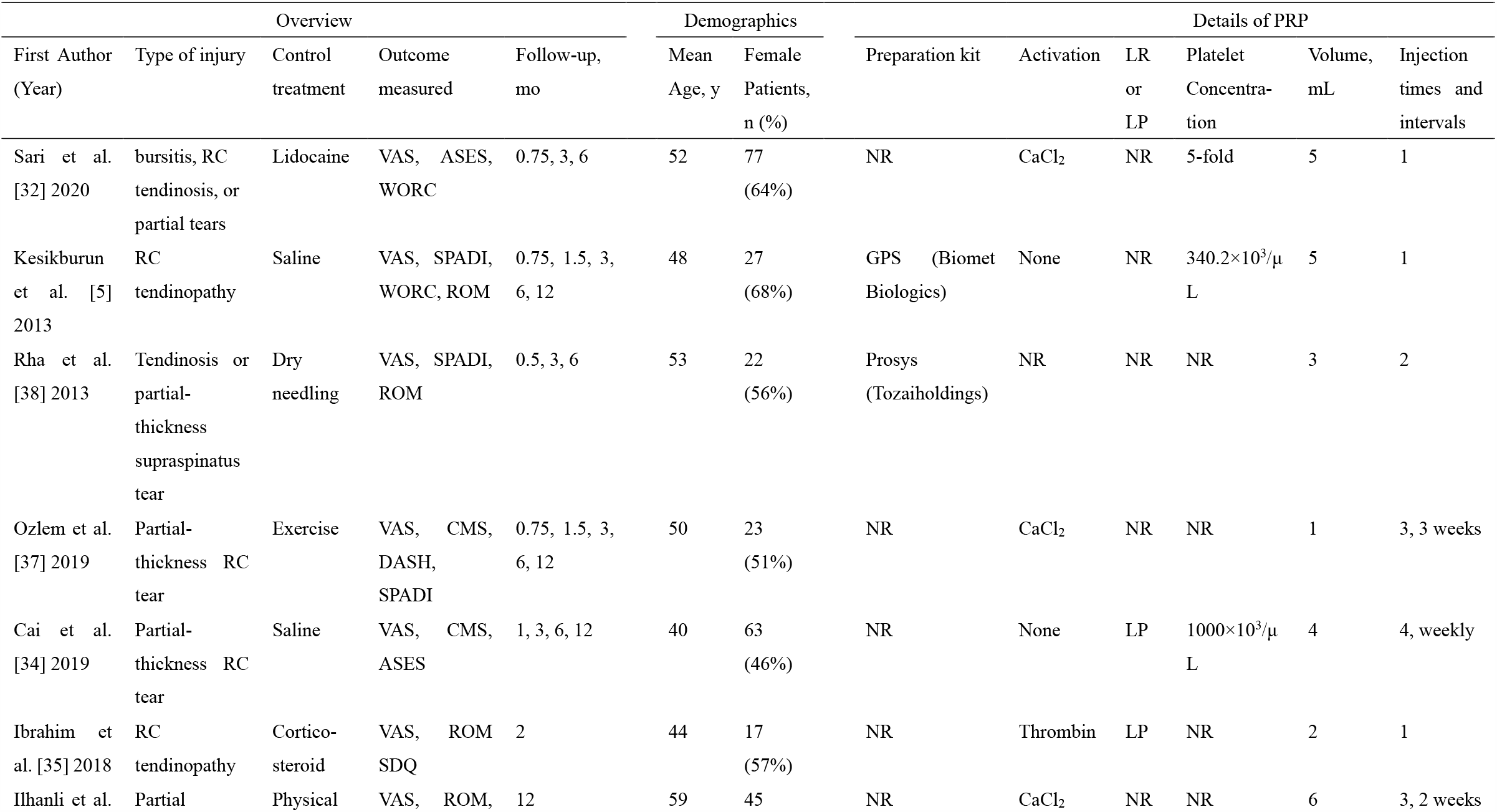
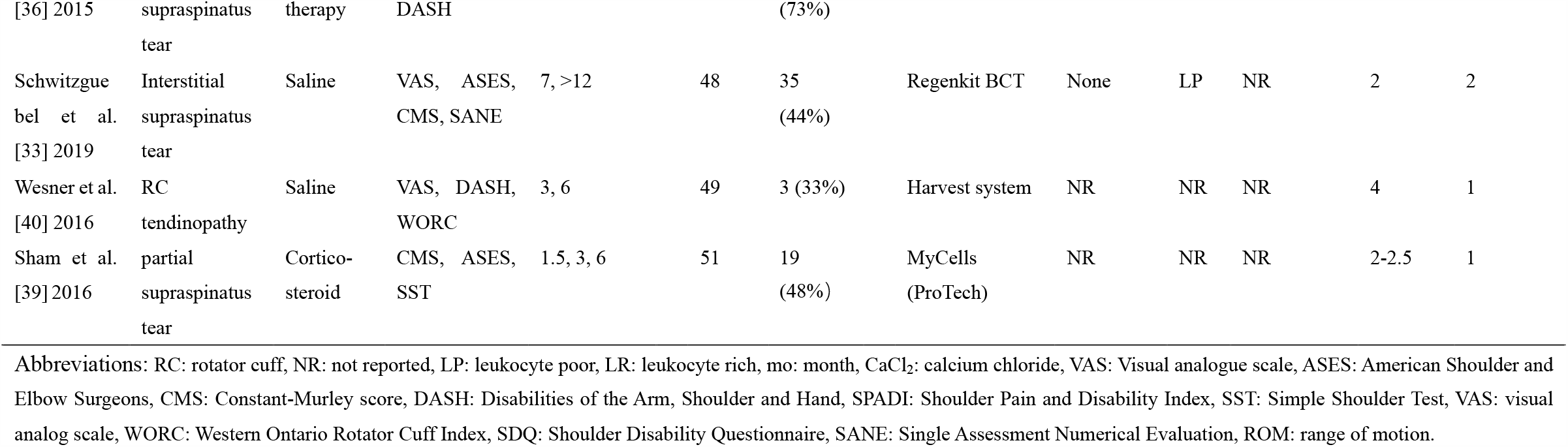
Characteristics of included Studies

### 3.2 Risk of bias in included studies

The results of the risk of bias assessment are presented in Figure 2. Among the ten studies, six studies were deemed to be at high risk of bias,^34-37, 39, 40^ two trials were deemed to be at low risk of bias,^5, 33^ and the remaining two studies were be at moderate risk.^32, 38^ Selection bias was the most common source of bias across the included studies. Adequate random sequence generation was reported in seven studies.^5, 32-34, 38-40^ Details of allocation concealment was reported in four studies.^5, 33, 37, 38^ Participants and personnel were blinded in five studies;^5, 32, 33, 38, 40^ and the blinding of outcome assessment was performed in seven studies.^5, 32-34, 36, 38, 40^ For attrition bias, a total of four studies had no description about incomplete outcome data, especially about reasons and statistical analysis for loss to follow-up.^32, 38-40^ There were significant differences in some of the baseline characteristics in Ilhanli et al.^36^ trial.

**Figure 2.**
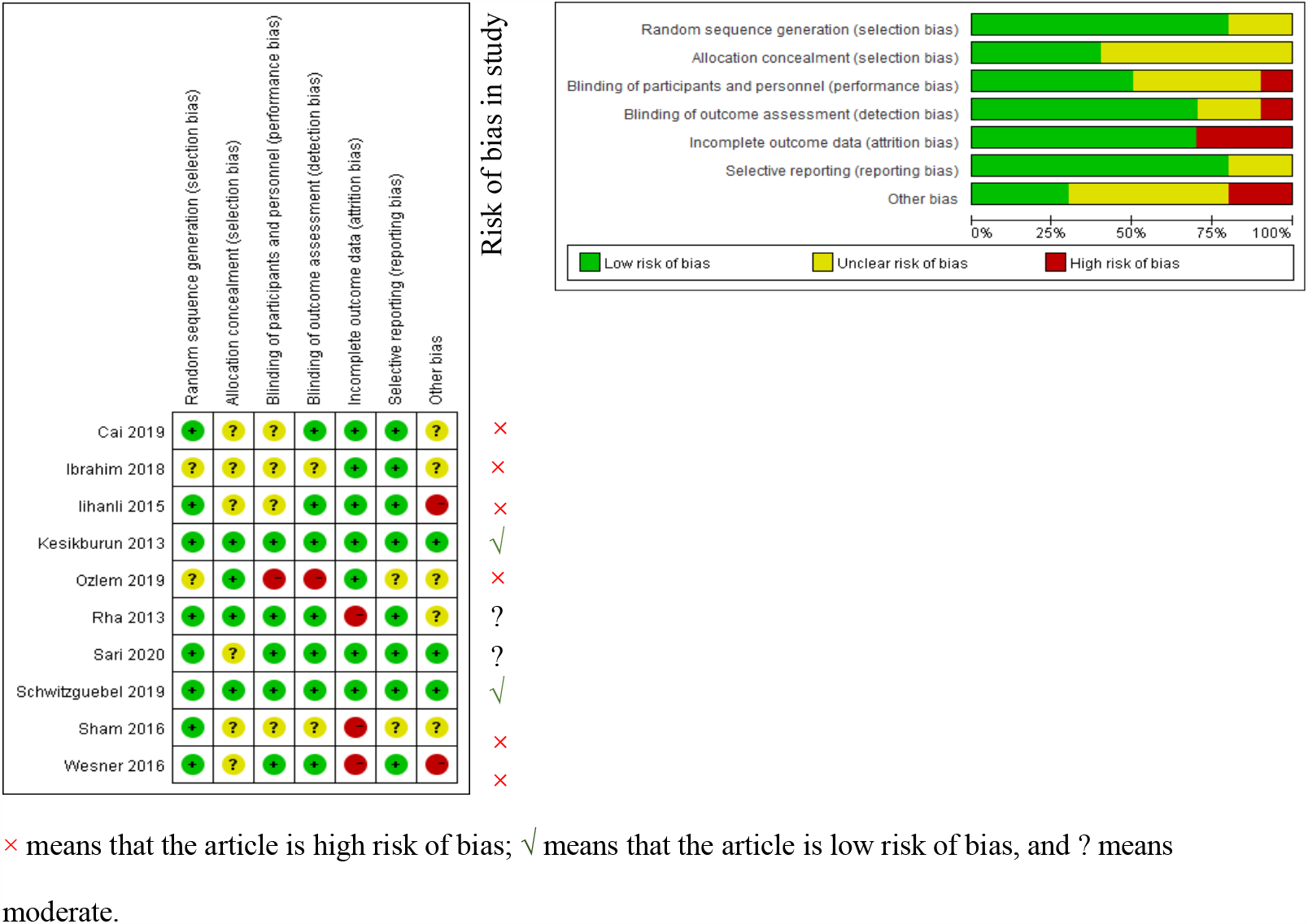
the graph shows quality assessment and risk of bias summary: review authors’ judgements about each risk of bias item for each included study by Cochrane Collaboration risk of bias tool.

### 3.3 Meta-analysis and sensitivity analysis

The results of the meta-analysis and sensitivity of nine studies is reported in Table 2.

**Table 2.**
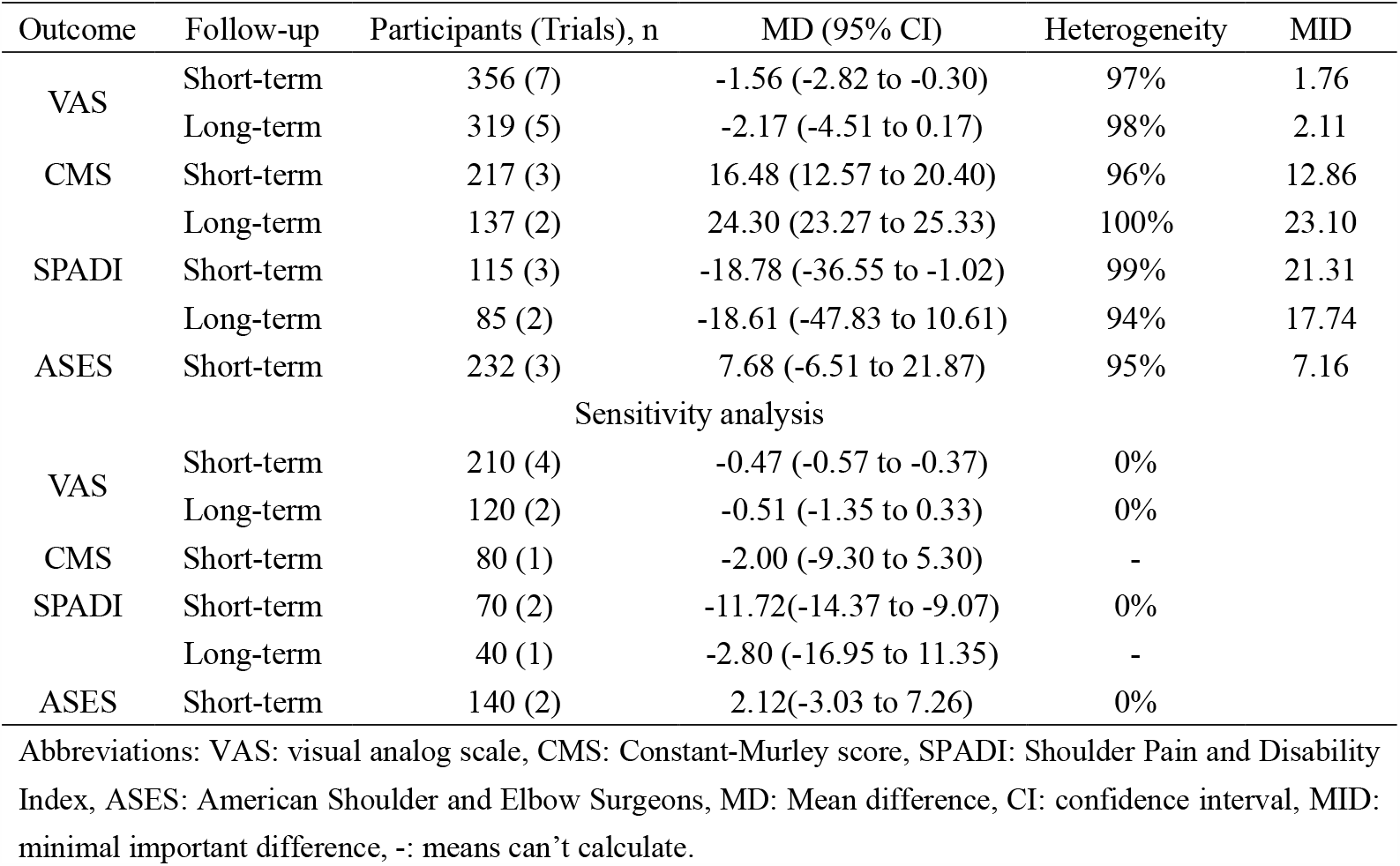
Summary of meta-analysis and sensitivity analysis for outcomes measures

#### 3.3.1 Pain

Outcomes of VAS are shown in the Figure 3. Patients who received PRP reported statistically significant improvements at short-term follow-up for VAS score (MD, - 1.56 [95% CI, -2.82 to -0.30]; *I*^2^=97%; *P*=.02). In order to assess the reliability of the results, studies with a high risk of bias were excluded from sensitivity analysis after removing 3 studies.^34, 37, 40^ Short-term VAS scores result was not altered (MD, -0.47 [-0.57 to -0.37]; *P*= .01) while the heterogeneity was substantially reduced (*I*^*2*^=0%). There were no statistical differences at long-term follow-up for VAS score (MD, -2.17 [95% CI, -4.51 to 0.17]; *I*^2^=98%; *P*= .007). Regarding the high heterogeneity for VAS, sensitivity analyses were also used to make results more reliable. After removing three studies^34, 36, 37^ with high risk of bias, the results were not altered (MD, -0.51 [95% CI, -1.35 to 0.33]; *I*^*2*^=0%; *P*= 0.74).

**Figure 3.**
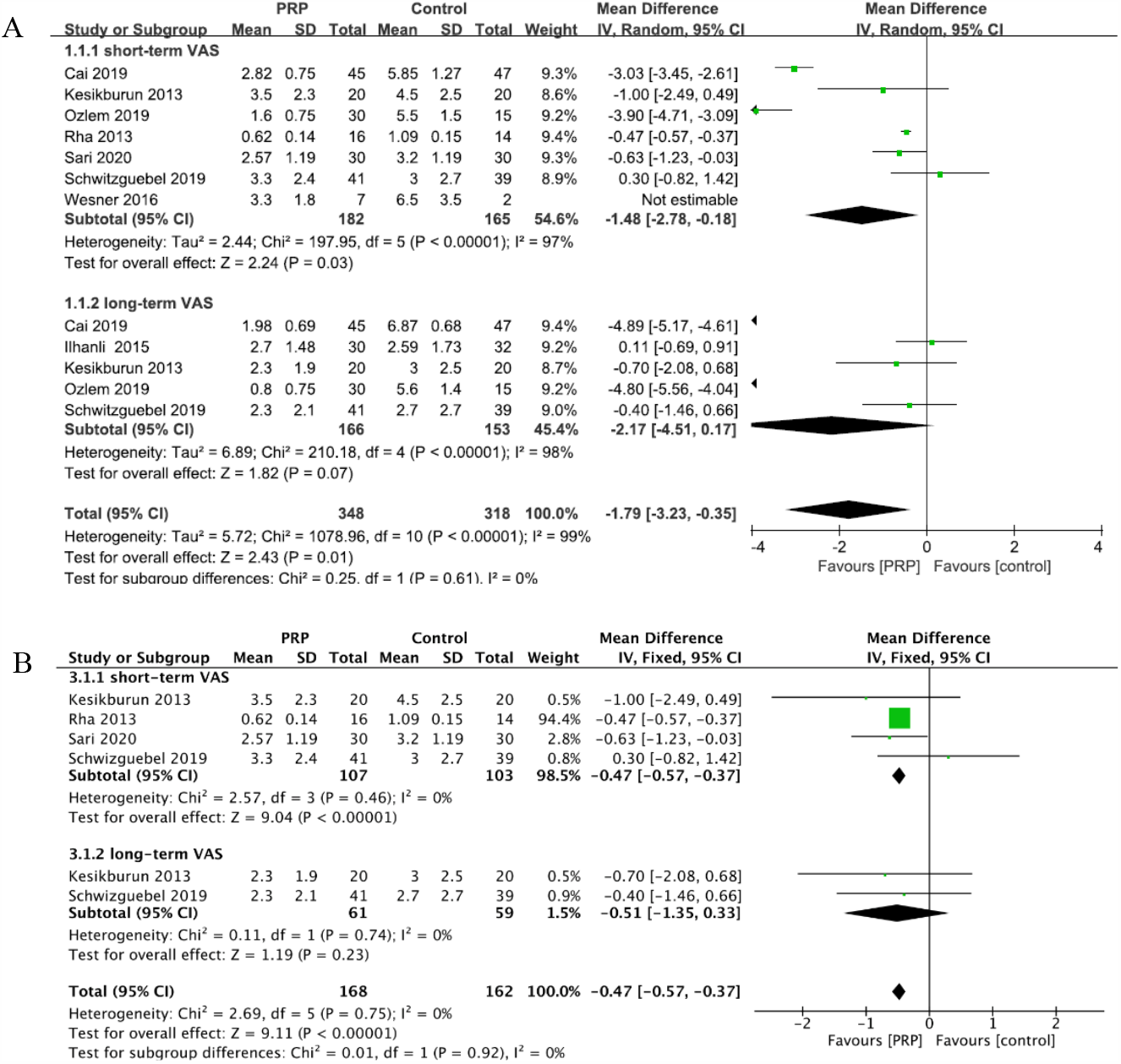
the graph shows forest plots of VAS score (A) and sensitivity analysis detailing mean differences (B). VAS: visual analogue scale.

#### 3.3.2 Function

The results of meta and sensitivity analysis of functional indexes are reported in Appendix A. Patients who received PRP showed statistically significant improvements at short-term follow-up for CMS (MD, 16.48 [95% CI,12.57 to 20.40]; *I*^2^=96%; *P*< .00001), and SPADI (MD, -18.78 [95% CI, -36.55 to -1.02]; *I*^2^=99%; *P*= .04), whereas the ASES score (MD, 7.68 [95% CI, -6.51 to 21.87]; *I*^2^=95%; *P*= .29). For SPADI, removing the study by Ozlem et al.^37^ did not alter conclusions (MD, -11.72 [-14.37 to -9.07]; *I*^*2*^=0%; *P*< .0001), while the heterogeneity was significantly decreased. For ASES scores, after removing 1 study^37^, heterogeneity was significantly decreased and result was not changed (MD, 2.12 [-3.03 to 7.26]; *I*^*2*^=0%; *P*= .36). Sensitivity analysis of CMS could not be performed after removing the Ozlem et al.^37^ and Cai et al.^34^

There were no statistical differences at long-term follow-up for SPADI (MD, - 18.61; 95% CI, -47.83 to10.61; *I*^2^ = 94%; *P* = .21). Additionally, there is no long-term ASES data to be pooled in this review. However, the CMS showed that PRP was statistical significantly more efficacious (MD, 24.30 [95% CI, 23.27 to 25.33]; *I*^2^=0%; *P*< .00001) based on data from two studies. Sensitivity analysis was unable to be performed due to lack of data after removing low quality studies.

### 3.4 Subgroup analysis

The following 3 factors were selected for subgroup analysis among the VAS scores and CMS: the PRP centrifugation approach (single or double), the number of PRP injections and rehabilitation after PRP injections. The forest plots of subgroup analysis are shown in Figure 4. The results of long-term CMS were not pooled due to insufficient data.

**Figure 4.**
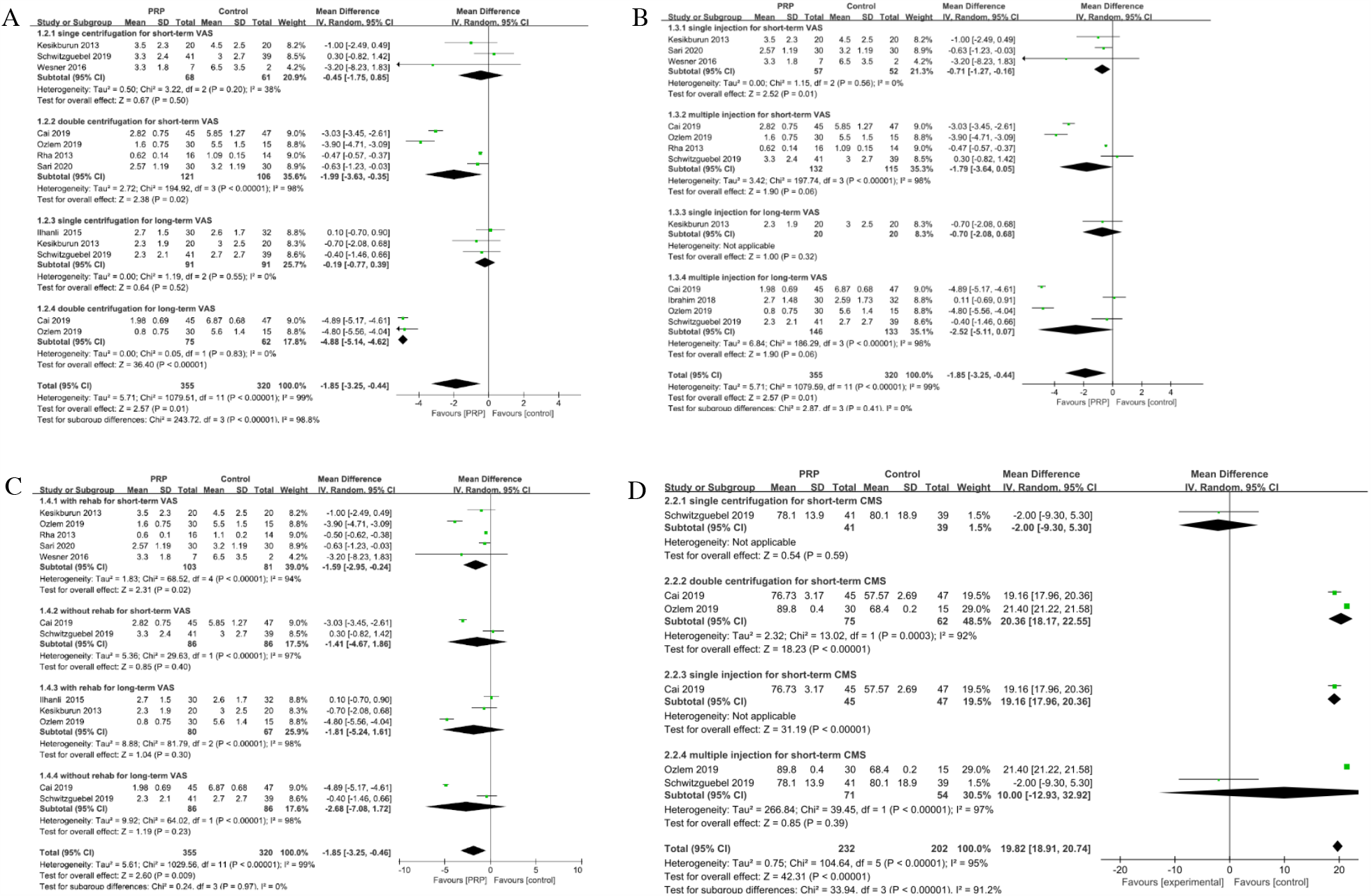
the graph shows VAS score based on the times of PRP centrifugation approach (A), number of injections (B), and followed rehabilitation (C) at short-term (6±1 months) and at long-term (≥1 year) follow-up. (D) shows short-term CMS based on centrifugation approach and number of injections. VAS: visual analogue scale, PRP: platelet-rich plasma, CMS: Constant-Murley score.

Patients who received PRP injections prepared by double centrifugation showed improvement in VAS score at short-term follow-up (MD, -1.99; 95% CI, -3.63 to -0.35; *I*^*2*^=98%; *P*= .02) and long-term follow-up (MD, -4.88; 95% CI, -5.14 to -4.62; *I*^*2*^=0%; *P*< .01). Moreover, CMS was improved at short-term follow-up in patients who received PRP injection prepared by double centrifugation (MD, 20.36; 95% CI, 18.17 to 22.55; *I*^*2*^=92%; *P*< .01). PRP prepared by single centrifugation did not appear to cause significant changes in VAS or CMS.

For the number of PRP injections subgroup, patients received the single injection appeared to reduce short-term VAS score (MD, -0.71; 95% CI, -1.27 to -0.16; *I*^*2*^=0%; *P*=.01). However, there was no significant difference in multiple PRP injections subgroup for short-term or long-term VAS scores (all *P*> .05) with substantial heterogeneity (*I*^2^=98%). There was no pooled evidence to demonstrate patients who received PRP with single injection had better CMS score at short-term. Additionally, there was no significant difference in CMS scores for multiple injection subgroup (MD, 10.00; 95% CI, -12.93 to 32.92; *I*^*2*^=97%; *P*=.39).

Among studies in which the inclusion/exclusion of post-injection rehabilitation was reported, patients who participated in PT had reduced VAS scores at short-term follow-up (MD, -1.59.00; 95% CI, -2.95 to 0.24; *P*=.02), while with significant heterogeneity (*I*^2^=94%). However, it appeared that long-term VAS score did not differ between compared groups. For studies with rehabilitation exclusion, there was no significant difference between treatment groups for short-term and long-term VAS scores (all *P*> .05).

## 4. Discussion

This study is the first level 1 meta-analysis to investigate the clinical efficacy of PRP in PTRCs and rotator cuff tendinopathy. The results of this study showed that PRP treatment reduced the pain and improved function (CMS and SPADI) statistically compared with the control groups at short-term follow-up. Nevertheless, no long-term improvements were observed for pain and function, except CMS score with a large heterogeneity.

Previous systematic review^16^ demonstrated that PRP injections may not have short-term benefit as a non-operative treatment of chronic rotator cuff disease but with limited evidence, in contrast to our findings. Nonetheless, Chen et al.^26^ reported that statistically improvements for functional outcomes in PRP-treated patients with rotator cuff tears comparing to controls, but none of them reached the MCIDs. Most of the controls in forementioned study were rotator cuff repair, which suggested PRP therapy was not worse than repair surgery. It is noteworthy that the MIDs of this study in pain scores between PRP and control groups were 1.76 at short-term and 2.11 at long-term follow-up. Both of them were beyond the MCID of pain (1.0). As for the secondary outcomes, the MIDs of the CMS and SPADI were all beyond their MCIDs. Nonetheless, short-term ASES (MID=7.16) did not reach its MCID (11.1). Hence, this suggests PRP injections provided statistically and clinically important improvements in pain relief and function for PTRCs and rotator cuff tendinopathy. However, the MCID seems to have obvious limitations. First, it is valid to be applied in groups but not the individuals. Second, it varies across different diseases, population and studies^41^. The conclusions of Chen et al. and ours both suggested potential benefits of PRP.

For subgroup analysis, the outcomes varied based on PRP centrifugation approach, number of PRP injections, and PRP alone or in combination with rehabilitation. Double centrifugation for PRP preparation was associated with lower VAS scores at short-term and long-term follow-up, as well as an increased CMS at short-term follow-up. The multiple number of PRP injections did not show any differences in the CMS and long-term VAS scores, while single injection of PRP resulted in statistically lower short-term VAS score. Rehab following the injections was associated with statistically better VAS scores in the short term.

PRP efficacy has been shown to be related to leukocyte concentration, platelet concentration and use of an exogenous activating agent, which can be modified via PRP preparation such as the speed of centrifugation, number of centrifugation cycles, and addition of products.^42^ In laboratory study showed that double centrifugation resulted in higher platelet concentration in rabbits, compared with a single centrifugation protocol.^43^ Additionally, there was higher platelet/leukocyte ratio on PRPs after second centrifugation according to de Vos RJ et al.^13^ Furthermore, the double centrifugation systems produce total white blood cell (WBC) counts at or above baseline, because they concentrate potentially beneficial monocyte or macrophage and lymphocyte WBC subpopulation.^44^ However, the double centrifugation technique was questioned because of the potential alterations in platelet morphology, which might affect functions.^45^ Other factors such as platelet concentration, the presence of WBC and activation agents were not included for this subgroup analyses, due to limited data. A simplistic definition of PRP is that the platelet count must be above baseline,^46, 47^ in addition to an increased platelet concentration.^48^ Besides this, the presence of WBC in PRP product has been a topic of considerable debate in the literature, regarding to the question of whether WBCs inhibit or promote tissue healing.^49^ Those inconsistent parameters of PRP may lead to the variability of the PRP efficacy and application among the published studies. Therefore, consensus recommendation by AAOS in 2018 and classification system for future biologic research by the American Academy of Physical Medicine and Rehabilitation both called for a standard PRP definition and preparation,^50, 51^ aiming to clarify the effect and reduce the heterogeneity among clinic trials.

There are several limitations to this study. The main limitation is the heterogeneity across these original studies, although sensitivity analysis was made to reduce the heterogeneity. First, the PRP preparation was inconsistent in all included RCTs and details of the kits, platelet concentration and leukocyte concentration were not fully-mentioned either^32-36^, which impeded further subgroups analysis. Although, we found that double centrifugation approach led to better results with relatively small number of studies. Second, the subsequent rehabilitation can be a source of co-intervention effect that may overestimate the results of PRP intervention. Third, in most studies we included, details of rotator cuff tear such as lesion sites (articular-sided, bursal-sided or intratendinous tears) or classification were missing, which may have contributed to the heterogeneity^52^. Furthermore, shoulder functional assessments varied across these studies, which included the DASH, CMS, Western Ontario Rotator Cuff Index, Single Assessment Numeric Evaluation, SPADI, ASES and quick DASH, raising difficulty to pool data in this meta-analysis. Another limitation is the inconsistent of control group interventions, consisting of exercise, physical therapy, dry needling and other injections.

## 5. Clinical messages

- PRP as a conservative treatment for PTRCs and rotator cuff tendinopathy.
- PRP-treated patients with PTRCs and rotator cuff tendinopathy demonstrated statistical and clinical improvements in pain and function within 6 months, although the effect may not last for a longer time.
- PRP with double centrifugation, single injection or rehabilitation followed could lead to better effects, although further studies were needed.

## Supporting information

Appendix. functional indexes

## Data Availability

Data have been provided in supplementary material.

## 6. Authors Contributions

XNX completed the searching, selected the included meta-analysis, appraised the quality, extracted data, and drafted the manuscript; JD completed the searching, selected the included meta-analysis, appraised the quality, extracted data, and help to drafting the manuscript; YL helped to judge the risk of bias; XY revised this manuscript; BC developed the protocol; HCH conceived of the study, and revised this manuscript. The all authors have read and approved the final version of the manuscript, and agree with the order of presentation of the authors.

