## Appendix. functional indexes for "Conservative treatment of partial-thickness rotator cuff tears and tendinopathy with platelet-rich plasma: a systematic review and meta-analysis"

**Appendix.** the graph shows forest plots of meta-analysis detailing mean differences comparing PRP injection with other conservative managements for CMS at short-term (6±1 months) and long-term (≥1 year) follow-up (A). (B) and (C) show the meta and sensitivity analysis of SPADI, respectively. (D) and (E) represent the meta and sensitivity analysis of ASES score, respectively. PRP: platelet-rich plasma, CMS: Constant-Murley score, SPADI: Shoulder Pain and Disability Index, ASES: American Shoulder and Elbow Surgeons.

A

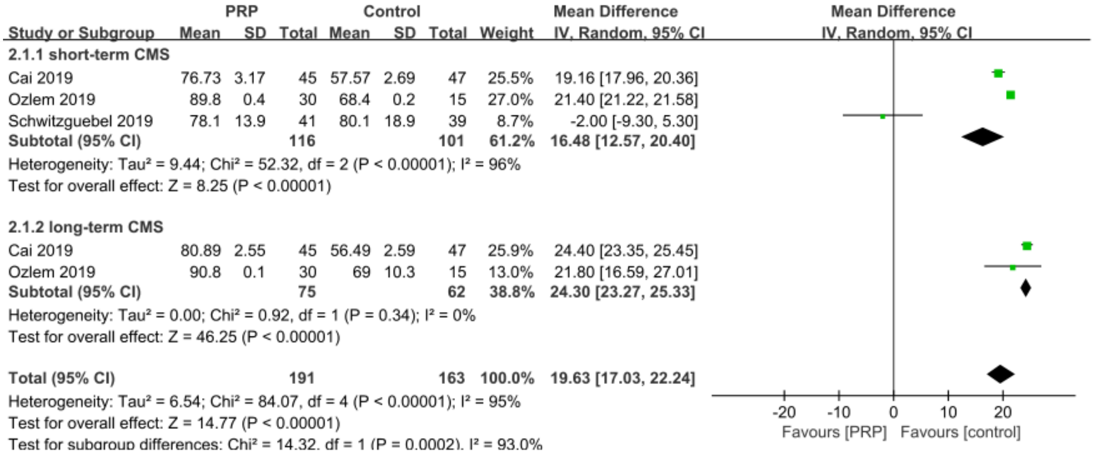


B

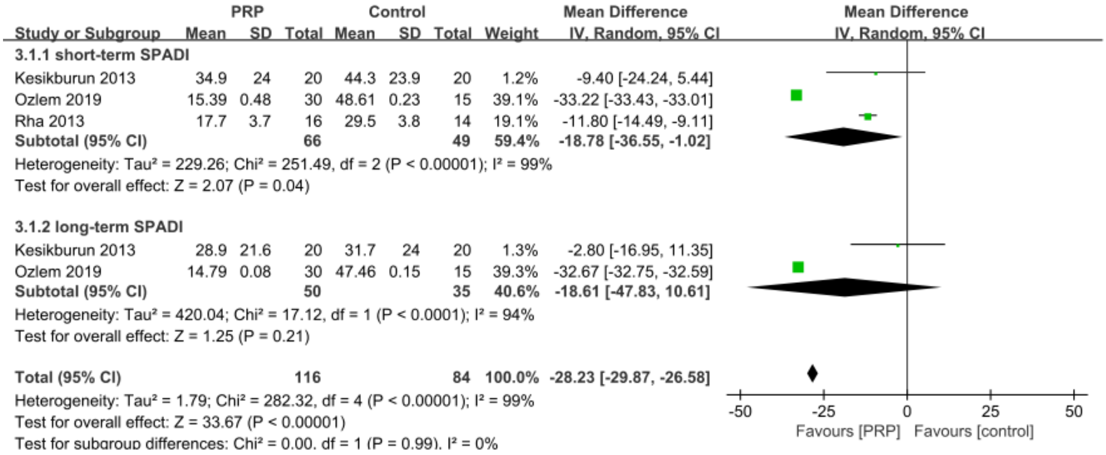


C

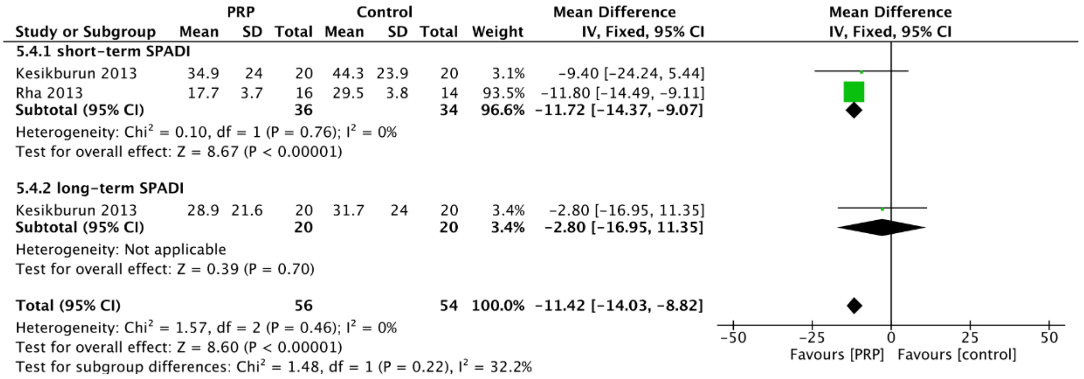

D

| Study or Subgroup | PRP Mean | PRP SD | PRP Total | Control Mean | Control SD | Control Total | Weight | Mean Difference IV, Random, 95% CI |
|---|---|---|---|---|---|---|---|---|
| **4.1.2 short-term ASES** | | | | | | | | |
| Cai 2019 | 67.89 | 5.76 | 45 | 48.21 | 2.52 | 47 | 35.7% | 19.68 [17.85, 21.51] |
| Sari 2020 | 63.87 | 11.96 | 30 | 60.27 | 11.92 | 30 | 33.7% | 3.60 [-2.44, 9.64] |
| Schwitzguebel 2019 | 70.2 | 19.7 | 41 | 72 | 24.7 | 39 | 30.6% | -1.80 [-11.62, 8.02] |
| **Subtotal (95% CI)** | | | **116** | | | **116** | **100.0%** | **7.68 [-6.51, 21.87]** |
| Heterogeneity: Tau² = 146.05; Chi² = 40.56, df = 2 (P < 0.00001); I² = 95% | | | | | | | | |
| Test for overall effect: Z = 1.06 (P = 0.29) | | | | | | | | |
| **Total (95% CI)** | | | **116** | | | **116** | **100.0%** | **7.68 [-6.51, 21.87]** |
| Heterogeneity: Tau² = 146.05; Chi² = 40.56, df = 2 (P < 0.00001); I² = 95% | | | | | | | | |
| Test for overall effect: Z = 1.06 (P = 0.29) | | | | | | | | |
| Test for subgroup differences: Not applicable | | | | | | | | |

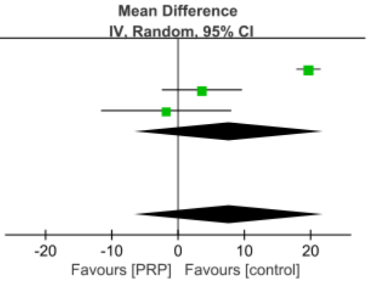


E

| Study or Subgroup | PRP Mean | PRP SD | PRP Total | Control Mean | Control SD | Control Total | Weight | Mean Difference IV, Fixed, 95% CI |
|---|---|---|---|---|---|---|---|---|
| **6.2.1 short-term ASES** | | | | | | | | |
| Sari 2020 | 63.87 | 11.96 | 30 | 60.27 | 11.92 | 30 | 72.5% | 3.60 [−2.44, 9.64] |
| Schwizguebel 2019 | 70.2 | 19.7 | 41 | 72 | 24.7 | 39 | 27.5% | −1.80 [−11.62, 8.02] |
| **Subtotal (95% CI)** | | | **71** | | | **69** | **100.0%** | **2.12 [−3.03, 7.26]** |
| Heterogeneity: Chi² = 0.84, df = 1 (P = 0.36); I² = 0% | | | | | | | | |
| Test for overall effect: Z = 0.81 (P = 0.42) | | | | | | | | |
| **Total (95% CI)** | | | **71** | | | **69** | **100.0%** | **2.12 [−3.03, 7.26]** |
| Heterogeneity: Chi² = 0.84, df = 1 (P = 0.36); I² = 0% | | | | | | | | |
| Test for overall effect: Z = 0.81 (P = 0.42) | | | | | | | | |
| Test for subgroup differences: Not applicable | | | | | | | | |

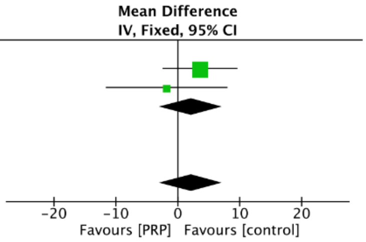
