## Supplementary Material for "Conservative treatment of partial-thickness rotator cuff tears and tendinopathy with platelet-rich plasma: a systematic review and meta-analysis"

### Search strategy

#### Medline (Ovid) search strategy

1. shoulder/
2. rotator cuff/
3. 1 or 2
4. exp bursitis
5. exp Rotator Cuff Injuries/
6. 4 or 5
7. 3 and 6
8. shoulder pain/
9. (Cuff Injury, Rotator or Injuries, Rotator Cuff or Injury, Rotator Cuff or Rotator Cuff Injury or Partial Thickness Rotator Cuff Tears or Incomplete Rotator Cuff Tear or Rotator Cuff Tears or Rotator Cuff Tear or Tear, Rotator Cuff or Tears, Rotator Cuff or Rotator Cuff Tendinosis or Rotator Cuff Tendinoses or Tendinoses, Rotator Cuff or Tendinosis, Rotator Cuff or Rotator Cuff Tendinitis or Rotator Cuff Tendinitides or Tendinitis, Rotator Cuff or Glenoid Labral Tears or Glenoid Labral Tear or Labral Tear, Glenoid or Labral Tears, Glenoid or Tear, Glenoid Labral).mp.
10. shoulder impingement syndrome/
11. rotator cuff injuries/
12. (rotator cuff or supraspinatus or infraspinatus or subscapular$ or teres).tw.
13. ((shoulder$ or subacromial or rotator cuff) adj5 (tendon$ or tendin$ or bursitis or calcium or calcif$ or impinge$ or tear$ or pain)).tw.
14. or/7-13
15. exp platelet rich plasma/
16. exp blood platelets/
17. (PRP or platelet rich plasma).mp.
18. exp Platelet derived growth factor/
19. platelet derived growth factor$.mp.
20. PDGF.mp.
21. exp Platelet Activation/

22. Platelet Activation.mp.
23. (platelet$ adj activat$).mp.
24. or/15-23
25. 14 and 24
26. Randomized controlled trial.pt.
27. Controlled clinical trial.pt.
28. (randomized or placebo or randomly or groups or trial).ab.
29. random*.ti,ab.
30. exp animals/ not humans.sh.
31. or/26-29
32. 31 not 30
33. 25 and 32

**Embase (Ovid) search strategy**

1. shoulder/
2. rotator cuff/
3. 1 or 2
4. calcium/
5. exp bursitis/
6. 4 or 5
7. 3 and 6
8. exp shoulder pain/
9. exp shoulder impingement syndrome/
10. exp rotator cuff/
11. ((shoulder* or rotator*) and (bursitis or frozen or impinge* or tendonitis or tendinitis or tendinopathy or pain*)).mp.
12. shoulder pain/
13. exp shoulder impingement syndrome/
14. exp rotator cuff injury/

15. (rotator cuff or supraspinatus or infraspinatus or subscapular$ or teres).tw.
16. ((shoulder$ or subacromial or rotator cuff) adj5 (tendon$ or tendin$ or bursitis or calcium or calcif$ or impinge$ or tear$ or pain)).tw.
17. 7 or 8 or 9 or 11 or 13 or 15 or 16
18. exp Platelet derived growth factor/
19. exp Thrombocyte/
20. exp Thrombocyte Rich Plasma/
21. (platelet$derived growth factor$ or PDGF).mp.
22. exp Thrombocyte Activation/
23. (platelet$rich plasma or PRP or platelet gel$).mp.
24. Platelet Activation.mp.
25. (platelet$ adj activat$).mp.
26. 18 or 19 or 20 or 21 or 22 or 23 or 25
27. 17 and 26
28. exp clinical trial/
29. exp evaluation studies/
30. (clin$ adj25 trial$).ti,ab. or ((singl$ or doubl$ or trebl$ or tripl$) adj25 (blind$ or mask$)).ti,ab. or (placebo$ or random$ or control$ or prospectiv$ or volunteer$).ti,ab.
31. (randomized controlled trial or randomization or double blind procedure or single blind procedure or methodology or follow up or prospective study or comparative study or placebo).sh.
32. animals/ not humans/
33. 28 or 29 or 30 or 31
34. 33 not 32
35. 34 and 27

**CENTRAL (Cochrane Library) search strategy**

1. shoulder/
2. rotator cuff/
3. exp bursitis/

4. shoulder pain/
5. (Cuff Injury, Rotator or Injuries, Rotator Cuff or Injury, Rotator Cuff or Rotator Cuff Injury or Rotator Cuff Tears or Rotator Cuff Tear or Tear, Rotator Cuff or Tears, Rotator Cuff or Rotator Cuff Tendinosis or Rotator Cuff Tendinoses or Tendinoses, Rotator Cuff or Tendinosis, Rotator Cuff or Rotator Cuff Tendinitis or Rotator Cuff Tendinitides or Tendinitis, Rotator Cuff or Glenoid Labral Tears or Glenoid Labral Tear or Labral Tear, Glenoid or Labral Tears, Glenoid or Tear, Glenoid Labral).mp.
6. shoulder impingement syndrome/
7. rotator cuff injuries/
8. (rotator cuff or supraspinatus or infraspinatus or subscapular$ or teres).tw.
9. ((shoulder$ or subacromial or rotator cuff) adj5 (tendon$ or tendin$ or bursitis or calcium or calcif$ or impinge$ or tear$ or pain)).tw.
10. or/1-9
11. exp platelet rich plasma/
12. exp blood platelets/
13. (PRP or platelet rich plasma).mp.
14. exp Platelet derived growth factor/
15. platelet derived growth factor$.mp.
16. PDGF.mp.
17. exp Platelet Activation/
18. Platelet Activation.mp.
19. (platelet$ adj activat$).mp.
20. or/11-19
21. 10 and 20 (126)
22. Randomized controlled trial.pt.
23. Controlled clinical trial.pt.
24. (randomized or placebo or randomly or groups or trial).ab.
25. random*.ti,ab.
26. exp animals/ not humans.sh.
27. or/22-25

28. 27 not 26

29. 28 and 21

**ISI Web of Science: Science Citation Index Expanded (SCI-EXPANDED) (1981 to January 2020), ISI Web of Science: Conference Proceedings Citation Index-Science (CPCI-S) (1982 to January 2020).**

1. TS=clinical trial*
2. TS=controlled trial*
3. TS=random*
4. TS=placebo*
5. #1 or #2 or #3 or #4
6. TS=platelet rich plasma
7. TS=blood platelets
8. TS=Platelet derived growth factor
9. TS=Platelet Activation
10. #6 OR #7 OR #8 OR #9
11. TS=Rotator Cuff Injuries
12. TS=shoulder pain
13. TS=Rotator Cuff Tendinosis
14. TS=shoulder impingement syndrome
15. #11 OR #12 OR #13 OR #14
16. #5 AND #10 AND #15

**Physiotherapy Evidence Database (PEDro)**

https://www.pedro.org.au/: platelet rich plasma.

http://www.opengrey.eu/search/request?q=greynet: platelet rich plasma.
